# SIGLEC1 enables straightforward assessment of type I interferon activity in inflammatory myopathies

**DOI:** 10.1101/2021.09.13.21263325

**Authors:** Manuel Graf, Sae Lim von Stuckrad, Akinori Uruha, Jens Klotsche, Lydia Zorn-Pauly, Nadine Unterwalder, Thomas Buttgereit, Martin Krusche, Christian Meisel, Gerd R. Burmester, Falk Hiepe, Robert Biesen, Tilman Kallinich, Werner Stenzel, Udo Schneider, Thomas Rose

**Author notes:** These authors contributed equally to this work (Udo Schneider, Thomas Rose). **Correspondence to:** Dr. med. Thomas Rose, Department of Rheumatology and Clinical Immunology, Charité – Universitätsmedizin Berlin, Charitéplatz 1, Berlin 10117, Germany.

## Abstract

**Objective:** To evaluate SIGLEC1 expression on monocytes by flow cytometry as a type I interferon biomarker in idiopathic inflammatory myopathies (IIM).

**Methods:** We performed a retrospective analysis of adult and pediatric patients with the diagnosis of IIM. SIGLEC1 expression was assessed by flow cytometry and was compared with Physician Global Assessment (PGA) or Childhood Myositis Assessment Scale (CMAS) disease activity scores. Mann Whitney-U test and receiver operating characteristic (ROC) curves were used for cross-sectional data analysis (n=96), two-level mixed-effects linear regression model for longitudinal analyses (n=26, 110 visits). Response to treatment was analyzed in 14 patients within 12 months, using Wilcoxon test. SIGLEC1 was compared to ISG15/MxA status by immunohistochemical staining of muscle biopsies (n=17).

**Results:** 96 patients with adult (a) and juvenile (j) dermatomyositis (DM, n=38), antisynthetase syndrome (AS, n=19), immune-mediated necrotizing myopathy (IMNM, n=8), inclusion body myositis (IBM, n=9), and overlap myositis (n=22) were included. SIGLEC1 distinguished significantly between active and inactive disease with an area under the curve (AUC) of 0.92 (95% CI: 0.83–1) in DM and correlated with disease activity longitudinally (aDM: standardized beta=0.54, p<0.001; jDM: standardized beta=-0.70, p<0.001). Response to treatment in DM was associated with a decreasing SIGLEC1 (p<0.01, Wilcoxon test). A positive ISG15/MxA stain was highly associated with a SIGLEC1 upregulation. SIGLEC1 was found upregulated in AS (42.1%) and IBM (22,2%) and not in IMNM.

**Conclusion:** SIGLEC1 is a candidate biomarker to assess type I interferon activity in IIM and proved useful for monitoring disease activity and response to treatment in juvenile and adult DM.

**Key messages:** *What is already known about this subject?:*

- There is an unmet need for routine clinical biomarkers to assess type I interferon activity in rheumatic musculoskeletal diseases
- SIGLEC1 is part of the type I interferon signature and transcripts were found to be upregulated in various autoimmune diseases such as systemic lupus erythematosus (SLE), Sjoegren syndrome and dermatomyositis.
- SIGLEC1 is expressed on the surface of monocytes and thus is easily assessable by flow cytometry, which enables straightforward monitoring of type I interferon activity

*What does this study add?:*

- An activation of the type I interferon system in IIM can be identified by flow cytometry analysing SIGLEC1 expression. SIGLEC1 correlated to disease activity and improvement under therapy in juvenile and adult dermatomyositis.

*How might this impact on clinical practice or future developments?:*

- SIGLEC1 expression is a suitable biomarker for monitoring type I interferon activation in rheumatic musculoskeletal diseases, which has clinical implications for patient stratification and treatment decision making especially in the context of interferon inhibitory therapies.

## Introduction

Idiopathic inflammatory myopathies are a group of autoimmune diseases that can affect the muscles, skin, lungs, joints and other organs. The EULAR/ACR classification of 2017 divides them into dermatomyositis, polymyositis, and inclusion body myositis.[1] However, increasing knowledge about sub-entities such as antisynthetase syndrome and immune-mediated necrotizing myopathy, and the discovery of new myositis-specific autoantibodies, has led to even more differentiation. Furthermore, the existence of polymyositis has recently been challenged.[2–4] Serum creatine kinase is used in clinical practice for diagnosis and monitoring of disease activity despite its shortcomings in subtypes of dermatomyositis.[5,6]

Studies have pointed out a crucial role of type I interferons in the etiopathogenesis of connective tissue diseases such as systemic lupus erythematosus and idiopathic inflammatory myopathies,[7,8] and therefore have become therapeutic targets of interest.[9–12] An upregulation of type I interferon-inducible transcripts was found in peripheral blood, muscle and skin biopsy specimens, particularly in patients with adult and juvenile dermatomyositis, showing a correlation with disease activity in adult and juvenile dermatomyositis.[7,13–20] However, type I interferon activation varies individually and is frequently not present. Thus an easy assessable type I interferon biomarker for patient stratification and disease activity monitoring in routine clinical diagnostics is highly needed.[21]

Analyzing the type I interferon signature in whole blood, SIGLEC1 (sialic acid binding Ig-like lectin 1, CD169), although monocyte-restricted, was found to be a highly upregulated transcript in patients with adult and juvenile dermatomyositis, systemic lupus erythematosus and various genetic interferonopathies.[13,22–25] However, the whole blood type I interferon signature is influenced by potentially disruptive factors (such as changes in immune blood cell distribution) that prevent optimal longitudinal comparative analysis.[26] Moreover, PCR testing and interpretation can be challenging and time-consuming. Hence, a cell-specific approach to analyzing type I interferon activity by measuring cell surface expression of SIGLEC1 on monocytes by flow cytometry could be advantageous.[26] The utility of SIGLEC1 assessment by flow cytometry has been shown very recently in juvenile dermatomyositis,[27] adult and juvenile systemic lupus erythematosus,[25,28–31] systemic sclerosis,[32] Sjögren’s syndrome,[33] genetic interferonopathies,[34] and viral infections including COVID-19,[35] but has not been analyzed in adult dermatomyositis and other subtypes of idiopathic inflammatory myopathies.

The aim of this study was to analyze the usefulness of flow cytometric measurement of SIGLEC1 expression on monocytes as a biomarker for type I interferon activity in patients with idiopathic inflammatory myopathies, and to determine if SIGLEC1 expression in blood correlates with disease activity, specific autoantibody profiles, response to treatment and type I interferon activity in skeletal muscle biopsies.

## Materials and Methods

### Standard Protocol Approvals, Registrations, and Patient consents

Prior approval for this study was obtained from the local ethics committee of the Charité - Universitätsmedizin Berlin (application number: EA2/094/19). Informed consent was not necessary for this retrospective study.

### Study population

Adult and Pediatric Rheumatology of Charité - Universitätsmedizin Berlin’s hospital electronic patient record system was searched (M.G., T.R) for patients with a diagnosis of dermatomyositis, polymyositis, antisynthetase syndrome, inclusion body myositis, immune-mediated necrotizing myopathy or overlap myositis and at least one flow cytometric measurement of SIGLEC1 expression on monocytes during the period between July 2015 and May 2020. Control groups consisted of patients with systemic lupus erythematosus and healthy individuals.

### Laboratory measurement of SIGLEC1 and autoantibodies

Blood samples for the analysis of SIGLEC1 (CD169) expression on monocytes, creatine kinase and autoantibody profiles were obtained on an outpatient or inpatient basis as part of routine diagnostics at the Charité - Universitätsmedizin Berlin, and they were analyzed at Labor Berlin – Charité Vivantes GmbH.

SIGLEC1 expression in whole blood was determined by flow cytometry using a highly standardized quantitative assay (Supplementary eText S1), as described by Stuckrad et al.[28] The lower limit of quantitation of this assay was 1200 monoclonal antibodies bound per cell (mAb/cell). The reference range for the expression of SIGLEC1 in healthy controls was determined to be less than 2400 mAb/cell.

Screening for IgG antinuclear antibodies (ANA) in human serum was performed using the AESKUSLIDES ANA-HEp-2 indirect immunofluorescence assay from AESKU.GROUP (Wendelsheim, Germany). Qualitative determination of ANA (SS-A 52, SS-A 60, SS-B, RNP-70, Sm, RNP/Sm, Scl-70, centromere B, and Jo1) in human serum was performed using the ANACombi ELISA from Orgentec (Mainz, Germany). Myositis-specific (anti-NXP2, anti-TIF1γ, anti-MDA5, anti-SRP, anti-Mi2, anti-OJ, anti-EJ, anti-PL7, anti-PL12, anti-Jo1, and anti-SAE) and myositis-associated autoantibodies (anti-Ku, anti-PM75, anti-PM100 and anti-Ro52) were determined in serum using the EUROLINE Autoimmune Inflammatory Myopathies 16 Ag line immunoassay (Euroimmun, Lübeck, Germany). Anti-HMGCR antibodies were determined using QUANTA Flash (Inova Diagnostics Inc., San Diego, CA, USA).

### Assessment of disease activity

Juvenile patients with idiopathic inflammatory myopathies were routinely evaluated with the Childhood Myositis Assessment Scale (CMAS) by a trained physiotherapist at the time of their visit, as described by Rider et al.[36] CMAS is a validated tool to measure muscle strength and endurance in juvenile idiopathic inflammatory myopathies.[36,37] Physiotherapists were not informed about laboratory parameters, such as creatine kinase or SIGLEC1 expression. The Physician Global Assessment (PGA), with scores ranging from 0 (no disease activity) to 10 (high disease activity), was used to rate overall disease activity in juveniles and adults. If PGA data was missing in the medical records, it was determined retrospectively by two experienced rheumatologists (U.S., A.v.S.) based on all available information except for SIGLEC1 expression. Visit 1 was defined as the date of the patient’s first SIGLEC1 expression measurement at our clinic.

### Response to treatment analysis

To assess a clinically meaningful response to treatment in juvenile and adult patients with dermatomyositis we included all patients with (a) active disease (PGA≥5) on the first visit (b) a follow up visit in a time frame of 3 to 12 months and (c) reduction of PGA by at least 20% (as proposed by Rider et al.[38]).

### Assessment of type I interferon activity in muscle biopsies

Interferon-stimulated gene 15 (ISG15) and myxovirus resistance protein A (MxA) reflect the type I interferon activity[39] and their protein expression can be assessed by immunohistochemical analysis. Briefly, 7-8 μm cryosections of skeletal muscle biopsies were stained using MxA (Santa Cruz polyclonal, Mx1/2/3, H-285, sc-50509, 1:100) and ISG15 (Abcam, clone ab14374, 1:50) as the primary antibodies, and immunohistochemical (IHC) analysis was performed using the iVIEW DAB (3,3’-diaminobenzidine) Detection Kit (Ventana Medical Systems, Tucson, Arizona, USA), as previously described.[40,41] Appropriate biotinylated secondary antibodies were used for signal amplification, and visualization of the reaction product was carried out on a Benchmark XT immunostainer (Ventana) using a standardized procedure. As previously described, MxA and ISG15 staining in the cytoplasm was considered positive except for necrotic or regenerating fibers. Fibers with equivocally faint staining were considered negative.[40,41] Biopsies were included in this study only if performed within seven days of SIGLEC1 measurement in blood.

### Statistical analysis

Quantitative data are presented as mean (range) or median (interquartile range), and qualitative data are presented as absolute numbers (percentage). The Mann Whitney U-test (MWU) was used to compare groups with non-normally distributed data. Spearman’s rank test was used to evaluate correlations between SIGLEC1 or creatine kinase and disease activity scores (PGA, CMAS). Receiver operating characteristic curves were plotted to compare the capability of SIGLEC1 and creatine kinase to distinguish between active (PGA≥5) and inactive disease (PGA<5).

Longitudinal data analyses were performed using a two-level mixed-effects linear regression model. Standardized beta coefficients (betaST) were reported to compare the strength of association between the following parameters of interest: (a) CMAS score and SIGLEC1 / creatine kinase, respectively, in juvenile dermatomyositis and (b) PGA score and SIGLEC1 / creatine kinase, respectively, in adult dermatomyositis. Clinically meaningful response to treatment was analyzed using Wilcoxon matched-pairs signed rank test. Linear mixed models analyses were performed with STATA 12.1, and Graphpad Prism Version 9.1.2 was used for all other data analyses and graphs. P values of <0.05 were considered statistically significant.

### Data availability

The datasets used and/or analyzed during the current study are available from the corresponding author on reasonable request.

## Results

### Study population

Seventy-four patients with confirmed diagnoses of idiopathic inflammatory myopathies were included (Table 1) and forty-four patients were excluded due to unclear diagnoses and/or insufficient clinical data. According to the EULAR/ACR criteria for idiopathic inflammatory myopathies,[1] 82.4% to 100% of the patients with adult and juvenile dermatomyositis, antisynthetase syndrome and inclusion body myositis had a probable or definite diagnosis, compared to only 62.5% of the immune-mediated necrotizing myopathy patients. All immune-mediated necrotizing myopathy diagnoses were conclusive and based on clinical and morphological criteria, as discussed at the 224th ENMC International Workshop.[42] The “Overlap” group consisted of twenty-two patients with overlap myositis (n=16), mixed connective tissue disease (n=5) and systemic sclerosis (n=1). No patient with isolated polymyositis as an independent entity could be identified in the time frame of 5 years.

**Table 1.** Patient characteristics. ^†^on first visit with assessment of SIGLEC1 ^‡^on first visit with assessment of SIGLEC1 and CMAS; NR, not reported; NA, not applicable; ND, not determined; IQR, interquartile range; n, absolute value; IIM, idiopathic inflammatory myopathies; DM, dermatomyositis; AS, antisynthetase syndrome; IMNM, immune-mediated necrotizing myopathy; IBM, inclusion body myositis; MCTD, mixed connective tissue disease; SLE, systemic lupus erythematosus; mAb, monoclonal antibody; CK, creatine kinase; CRP, c-reactive protein; JAK, Janus Kinase; MMF, Mycophenolate Mofetil; CsA, Ciclosporin A; Cyc, Cyclophosphamide; AZA, Azathioprine; EULAR, European League Against Rheumatism; ACR, American College of Rheumatology PGA, physician global assessment; CMAS, Childhood Myositis Assessment Scale; SLEDAI-2k, Systemic Lupus Erythematosus Disease Activity Index 2000

| Patient characteristics | Adult DM | Juvenile DM | AS | IMNM<br>(adult +<br>juvenile) | IBM | Control 1:<br>Overlap (adult<br>+ juvenile) | Control 2:<br>SLE | Control 3:<br>Healthy<br>individuals |
| --- | --- | --- | --- | --- | --- | --- | --- | --- |
|  | (n=21) | (n=17) | (n=19) | (n=8) | (n=9) | (n=22) | (n=30) | (n=19) |
| Female, n (%) | 10 (47.6%) | 9 (53%) | 10 (52.6%) | 3 (37.5%) | 6 (66.6%) | 20 (90.9%) | 26 (86.67%) | 10 (52.63%) |
| Age at diagnosis, years, mean (range) | 52.3 (20-84) | 9 (2-16) | 53.3 (17-77) | 56.9 (6-86) | 67 (52-79) | 39.5 (10-77) | 41.7 (19-70) | ND (18-62) |
| Time since first diagnosis <sup>†</sup>, n (%) |  |  |  |  |  |  |  |  |
| <3 months | 10 (47.6%) | 7 (41.2%) | 6 (31.6%) | 4 (50%) | 0 | 8 (36.4%) | 14 (46.7%) | NA |
| 3 to 12 months | 3 (14.3%) | 2 (11.8%) | 5 (26.3%) | 2 (25%) | 1 (11%) | 3 (13.7 %) | 3 (10%) | NA |
| >12 months | 8 (38%) | 8 (47.1%) | 8 (42.1%) | 2 (25%) | 8 (89%) | 11 (50%) | 13 (43.3%) | NA |
| SIGLEC1 expression, mAb/cell,<br>median (IQR) <sup>†</sup> | 5876 (1211-10282) | 5272 (1200-12691) | 1580 (1200-3390) | 1246 (1200-1853) | 1949 (1306-4602) | 1661 (1200-6915) | 7947 (3695-11924) | 1200 (1200-1200) |
| CK, U/l, median (IQR) <sup>†</sup> | 109 (74.5-442.5),<br>NR=1 | 218.5 (116.3-1988), NR=1 | 627 (102.3-2173), NR=1 | 2118 (445.8-4353) | 497 (322.5-988.5),<br>NR=1 | 421 (68-962.3),<br>NR=4 | 62.5 (45.4-97.3) | ND |
| CRP, mg/l, median (IQR) <sup>†</sup> | 4.95 (0.7-24),<br>NR=1 | 0.8 (0.3-3.3),<br>NR=2 | 5.5 (1.9-29.1),<br>NR=1 | 2.75 (0.68-11.3),<br>NR=2 | 3.3 (1.3-33),<br>NR=1 | 3.0 (0.9-7.4), NR=2 | 2.4 (0.7-17.3) | ND |
| Biopsy-confirmed IIM diagnosis /<br>performed biopsies | 14/15 | 6/7 | 7/9 | 6/7 | 9/9 | 0/10 | NA | NA |
| “Definite” or “probable” IIM as per<br>EULAR/ACR criteria, n (%) | 19 (90.5%) | 13 (82.4%) | 17 (89.5%) | 5 (62.6%) | 9 (100%) | NA | NA | NA |
| PGA, mean (range) <sup>†</sup> | 6 (2-10) | 4.4 (0-8) | 5.4 (2-9) | 4.9 (2-8) | 4.22 (3-6) | 4.5 (3-9), NR=7 | NA | NA |
| CMAS, mean (range) <sup>‡</sup> | NA | 35 (2-52),<br>NR=4 | NA | NA | NA | NA | NA | NA |
| SLEDAI-2k, mean (range) <sup>†</sup> | NA | NA | NA | NA | NA | NA | 6.6 (0-24) | NA |
| Prednisolone, number of patients (mean in mg/day on visit 1) | 12 (36.1 mg/day) | 9 (8.3 mg/day), NR=1 | 16 (13.3 mg/day) | 4 (13.1 mg/day) | 3 (7.5 mg/day) | 9 (6.9 mg/day), NR=1 | 17 (NR) | NA |
| Other medication <sup>†</sup>, n (%) |  |  |  |  |  |  |  |  |
| Hydroxychloroquine/Chloroquine | 1 (4.8%) | 2 (11.8%) | 3 (15.8%) | - | - | 3 (13.7%) | 10 (33.3%) | NA |
| Intravenous Immunoglobulin | 2 (9.5%) | 2 (11.8%) | 1 (5.3%) | 1 (12.5%) | 1 (11.1%) | - | - | NA |
| Methotrexate | 1 (4.8%) | 6 (35.3%) | 3 (15.8%) | 1 (12.5%) | 1 (11.1%) | 4 (18.2%) | 1 (3.3%) | NA |
| Rituximab | - | - | 2 (10.5%) | - | 1 (11.1%) | - | - | NA |
| JAK Inhibitor | 3 (14.3%) | - | - | - | - | - | - | NA |
| MMF / CsA / Cyc / AZA | 5 (23.8%) | 3 (17.6%) | 5 (26.3%) | 2 (25%) | - | 3 (13.7%) | 6 (20%) | NA |
| No medication | 8 (38.1%) | 7 (41.2%) | 3 (15.8%) | 4 (50%) | 5 (55.6%) | 10 (45.5%) | 13 (43.3%) | 19 (100%) |

### SIGLEC1 expression in inflammatory myopathies and control groups

Adult and juvenile dermatomyositis patients expressed high levels of SIGLEC1 (median, 5876 and 5272 mAb/cell) (Figure 1 and Table 1). SIGLEC1 was upregulated in all newly diagnosed juvenile dermatomyositis patients (n=4, median, 13735 mAb/cell) and in 77.8% (n=7) of the newly diagnosed adult dermatomyositis patients (n=9, median, 9812 mAb/cell). There was no significant difference in SIGLEC1 expression between adult/juvenile dermatomyositis and systemic lupus erythematosus (systemic lupus erythematosus vs. adult dermatomyositis: p=0.172; systemic lupus erythematosus vs. juvenile dermatomyositis: p=0.485; MWU). Elevated SIGLEC1 expression was observed in two patients with inclusion body myositis (both anti-Ro52^+^, one anti-U1RNP^+^ and anti-Ku^+^). In the overlap group, six patients showed elevated SIGLEC1 levels: four with mixed connective tissue disease (anti-U1RNP^+^), one with anti-Ku antibodies, and one with overlap to juvenile systemic lupus erythematosus. In patients with systemic lupus erythematosus, SIGLEC1 correlated with the systemic lupus erythematosus disease activity index (SLEDAI-2k) (r=0.46, p=0.013, Spearman’s r).

**Figure 1.**
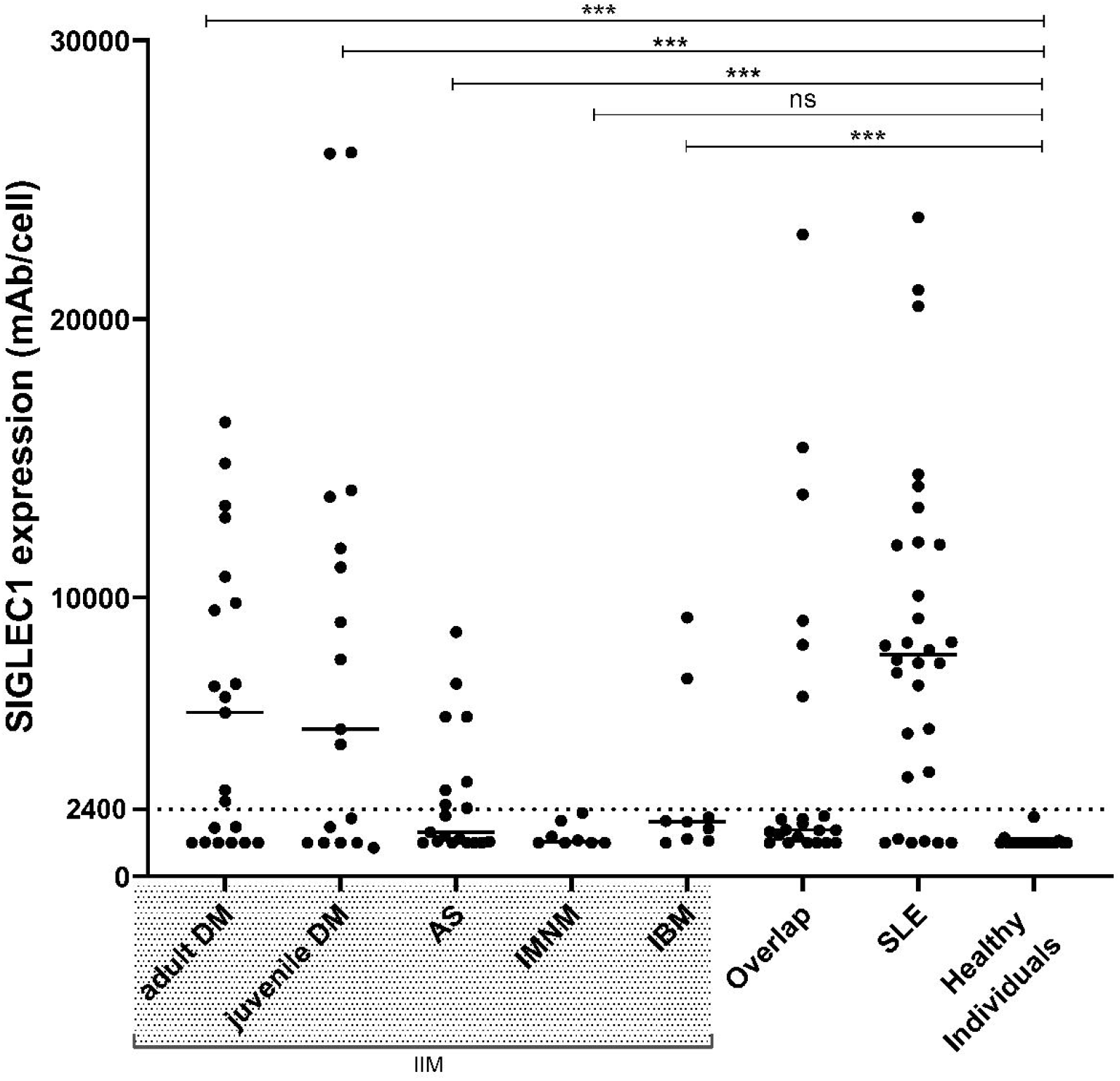
SIGLEC1 expression on monocytes of all patients at first visit. Horizontal bars show median values. Mann-Whitney-U-test was used to compare patients with the following idiopathic inflammatory myopathies (IIM) — adult (n=21) and juvenile (n=17) dermatomyositis (DM), antisynthetase syndrome (AS, n=18), immune-mediated necrotizing myopathy (IMNM, n=8), inclusion body myositis (IBM, n=9) — with healthy individuals (n=19); ***, p<0.001; ns, not significant; SLE, systemic lupus erythematosus

### SIGLEC1 expression and disease activity

To determine if SIGLEC1 expression is associated with disease activity, each subgroup of idiopathic inflammatory myopathies was divided into two groups by PGA score: PGA<5 (no to moderate disease activity) and PGA≥5 (moderate to severe disease activity) (Figure 2 and Figure 3). There was a significant difference between the two groups in adult dermatomyositis (p<0.001, MWU) and juvenile dermatomyositis (p<0.001, MWU). SIGLEC1 levels generally correlated with PGA levels in adult and juvenile dermatomyositis (adult dermatomyositis: r=0.81, p<0.001 and juvenile dermatomyositis: r=0.80, p<0.001; Spearman’s r) (Supplementary eFigure S3). We also analyzed the capability of SIGLEC1 and creatine kinase to distinguish between PGA<5 and PGA≥5 and found a high area under the curve (AUC) for SIGLEC1 in dermatomyositis (AUC=0.92, p<0.001) and both dermatomyositis subgroups (juvenile dermatomyositis: AUC=0.97, p=0.001; adult dermatomyositis: AUC=0.96, p=0.002). The AUC was lower for creatine kinase (dermatomyositis: AUC=0.71; p=0.04; juvenile dermatomyositis: AUC=0.94, p=0.003; adult dermatomyositis: AUC=0.60, p=0.513) (Figure 2 and Supplementary eFigure S1). To identify the best cut-off point for SIGLEC1 in dermatomyositis, we applied the Youden-index (J = sensitivity + specificity – 1) and found a lower threshold of 2383 mAb/monocyte (sensitivity 87,5%; specificity 85,71%), with a positive predictive value of 91.3%. In juvenile dermatomyositis, SIGLEC1 correlated with CMAS (r=-0.57, p=0.046; Spearman’s r) in cross-sectional analysis, while creatine kinase showed no significant correlation with CMAS (r=-0.40, p=0.180, Spearman’s r) (Supplementary eFigure S2).

**Figure 2.**
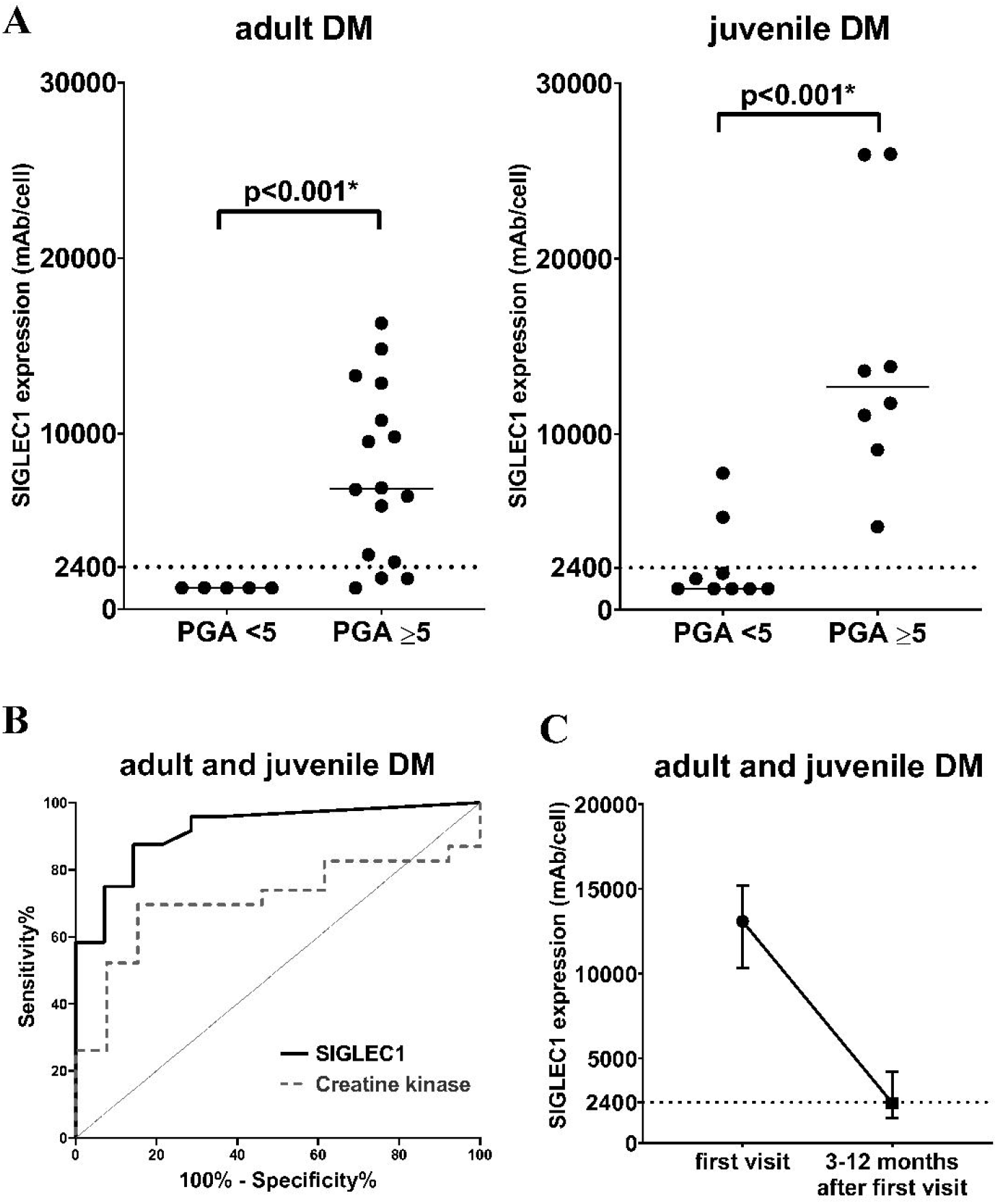
SIGLEC1 expression in DM patients. **(A)** adult and juvenile DM subgroups (n=21/n=17) are separated by PGA score: PGA<5 (no to moderate disease activity) and PGA≥5 (moderate to severe disease activity). Horizontal bars show median values; asterisks (*) represent significant results (p<0.05). The Mann-Whitney test was used to compare groups. PGA, Physician Global Assessment **(B)** Receiver operating characteristic curves for SIGLEC1 and creatine kinase in juvenile and adult DM (n=38). The curves show the ability of each biomarker to distinguish between patients with PGA≥5 (moderate to severe disease activity) and PGA<5 (no to moderate disease activity): SIGLEC1: AUC=0.92, 95%CI 0.83-1; p<0.001; CK: AUC=0.71; 95%CI 0.54-0.89; p=0.04 **(C)** SIGLEC1 expression in adult and juvenile DM patients with a clinically meaningful improvement between the first visit and 3-12 months after the first visit (n=14, p<0.01, Wilcoxon test). The median and interquartile range for each time point is shown.

**Figure 3.**
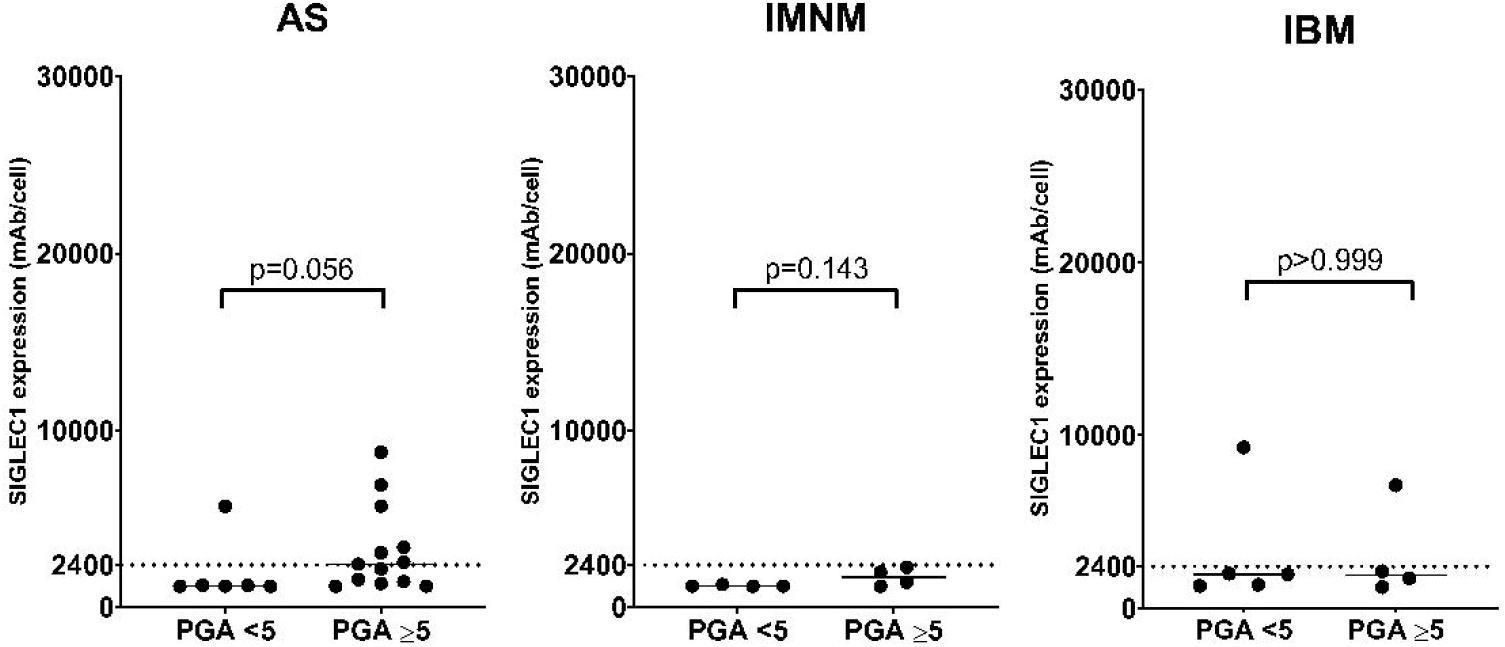
SIGLEC1 expression versus disease activity in AS, IMNM and IBM subgroups, separated by PGA score: PGA<5 (no to moderate disease activity) and PGA≥5 (moderate to severe disease activity). Horizontal bars show median values. The Mann-Whitney test was used to compare groups. PGA, Physician Global Assessment

In the longitudinal analyses based on a total of 65 visits by 12 juvenile dermatomyositis patients (SIGLEC1 vs. CMAS) and 45 visits by 14 adult dermatomyositis patients (SIGLEC1 vs. PGA), SIGLEC1 correlated with disease activity in juvenile (SIGLEC1 vs. CMAS: betaST=-0.70, p<0.001) and adult dermatomyositis (SIGLEC1 vs. PGA: betaST=0.54, p<0.001) (Table 2). The correlations between disease activity and SIGLEC1 were stronger than those between disease activity and creatine kinase in all analyses. The increase or decrease of SIGLEC1 between two consecutive visits was associated with changes in disease activity scores. Results for creatine kinase were not significant. Longitudinal graphs of biomarkers and disease activity are presented in Supplementary eFigure S4 and S5.

**Table 2.**
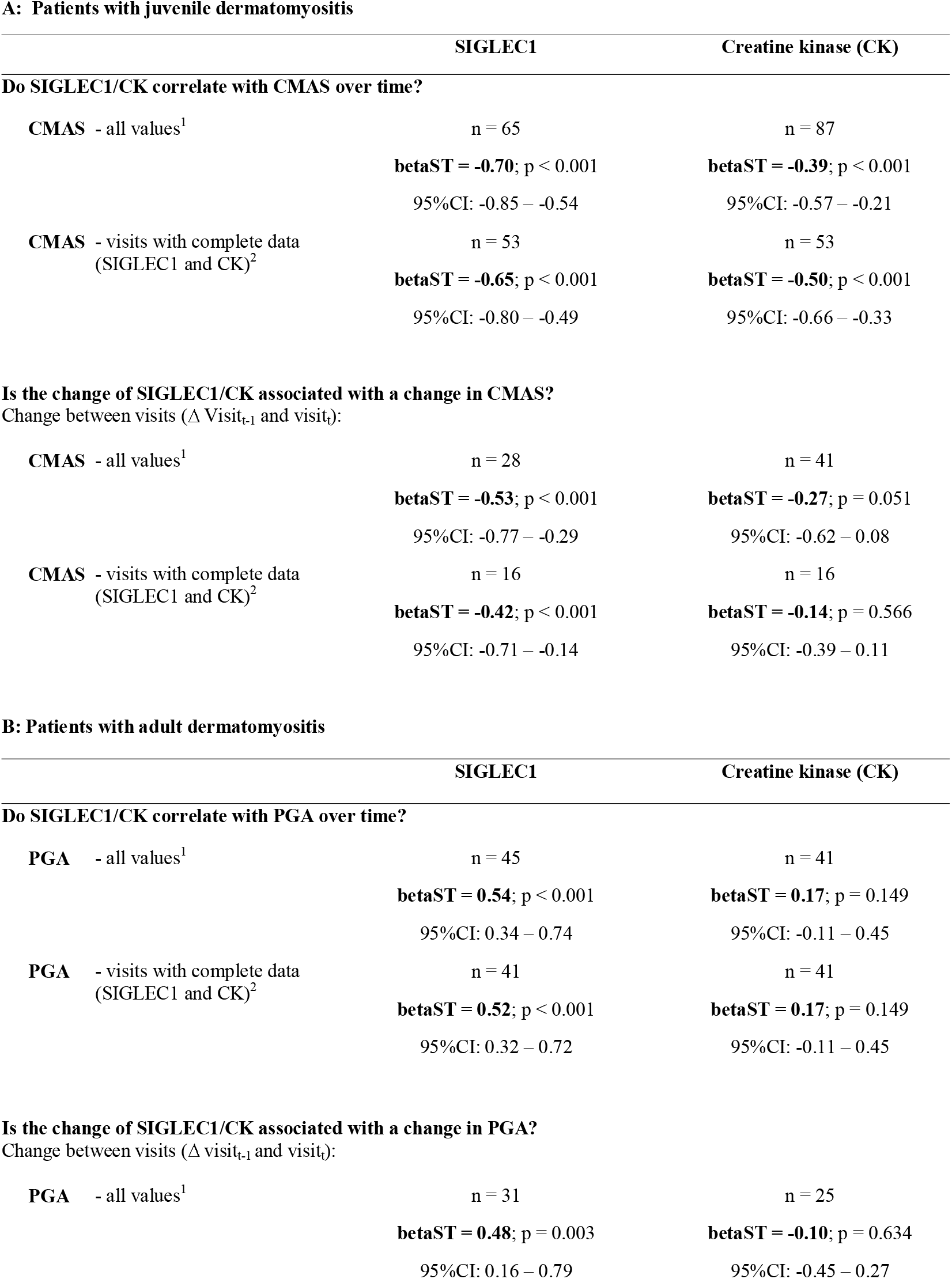
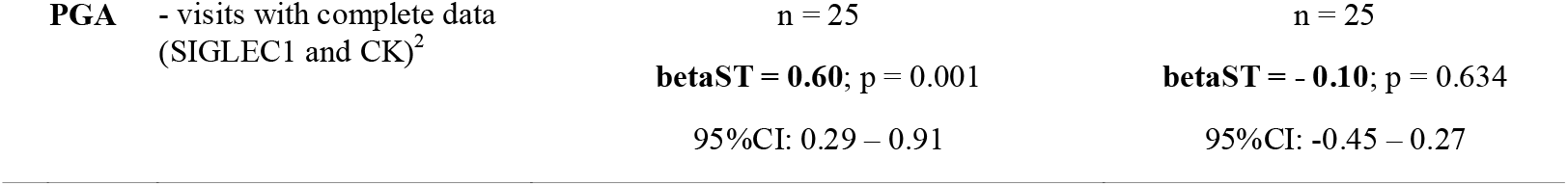
Results of longitudinal analyses comparing SIGLEC1 and creatine kinase (CK) with disease activity scores (CMAS/PGA) for (A) 12 juvenile and (B) 14 adult dermatomyositis patients. Statistical analysis was performed using a two-level mixed-effects linear regression model. Abbreviations: betaST, standardized beta coefficient; n, number of analyzed values; CI, confidence interval; PGA, Physician Global Assessment; CMAS, Childhood Myositis Assessment Scale; CK, Creatine Kinase; ^1^ SIGLEC1 and CK values were analyzed independently of each other. ^2^ Included only those visits where both biomarkers (SIGLEC1 and CK) were assessed (complete case analysis)

14 patients with dermatomyositis (adult, n=6 and juvenile, n=8) fulfilled the inclusion criteria for our response to treatment analysis. Between the two visits, there was a significant reduction of SIGLEC1 expression (median of differences -10059, IQR -6058 to -12152, p<0.01, Wilcoxon test) (Figure 2). PGA scores between the respective visits improved by -76.5% (median of differences, IQR -63% to -91%). Medication for the treatment were as followed: Prednisolone (n=14), Methotrexate (n=9), Intravenous Immunoglobulin (n=9), Hydroxychloroquine (n=4), Azathioprine (n=3), Cyclophosphamide (n=2).

No significant correlation between SIGLEC1 and disease activity was detected in patients with antisynthetase syndrome, inclusion body myositis or immune-mediated necrotizing myopathy (Figure 3 and Supplementary eFigure S3).

### SIGLEC1 expression and myositis-specific autoantibodies

Subgroup analysis of juvenile and adult dermatomyositis patients with moderate to high disease activity (PGA≥5) revealed elevated SIGLEC1 expression in 5/5 patients with TIF1γ-antibodies, 6/7 with MDA5-antibodies, 4/5 with NXP2-antibodies, and 2/3 with Mi2-antibodies. Those with immune-mediated necrotizing myopathy-associated antibodies (anti-SRP, anti-HMGCR) had consistently low SIGLEC1 expression (Figure 4).

**Figure 4.**
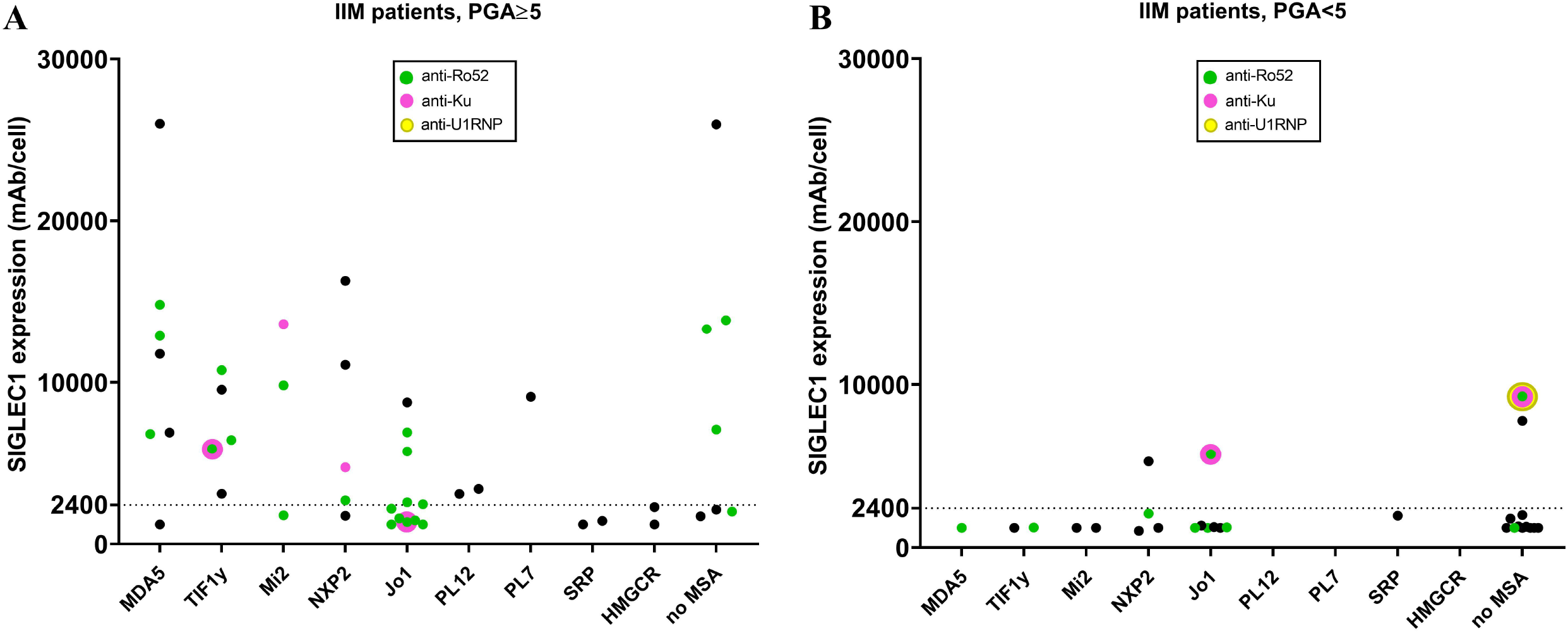
SIGLEC1 expression and autoantibody status. SIGLEC1 expression of all IIM patients (n=74) at first visit. Each group represents patients with a positivity for the respective myositis specific autoantibody (MSA) and (A) PGA≥5 (moderate to severe disease activity) and (B) PGA<5 (no to moderate disease activity); if present, additional myositis associated antibodies (MAA) are marked by different colors and no MAA is resembled by black.

### Comparison of SIGLEC1 expression on monocytes with ISG15/MxA status in muscle tissue

Of the 17 muscle samples eligible for immunohistochemical staining (inclusion criteria: 7 days or less between SIGLEC1 measurement and muscle biopsy), 7 (41.2%) were positive for ISG15 and/or MxA. The average time between SIGLEC1-measurement and muscle biopsy was 3 days (range: 0-7 days). All muscle biopsy patients that were positive for ISG15/MxA (in staining) had elevated SIGLEC1 expression on monocytes in peripheral blood (Figure 5), but one patient with antisynthetase syndrome had only minimal elevation (2449 mAb/cell). Three patients with upregulation of SIGLEC1 expression had a negative ISG15/MxA status and were diagnosed with dermatomyositis: one had anti-TIF1γ antibodies, one did not have any myositis-specific autoantibodies, and one had anti-MDA5^+^ amyopathic dermatomyositis. One patient with inclusion body myositis who was positive for SIGLEC1 expression on monocytes and in immunohistochemical staining in muscle biopsy was also positive for anti-Ku, anti-U1RNP and anti-Ro52 antibodies (Supplementary eTable S1).

**Figure 5.**
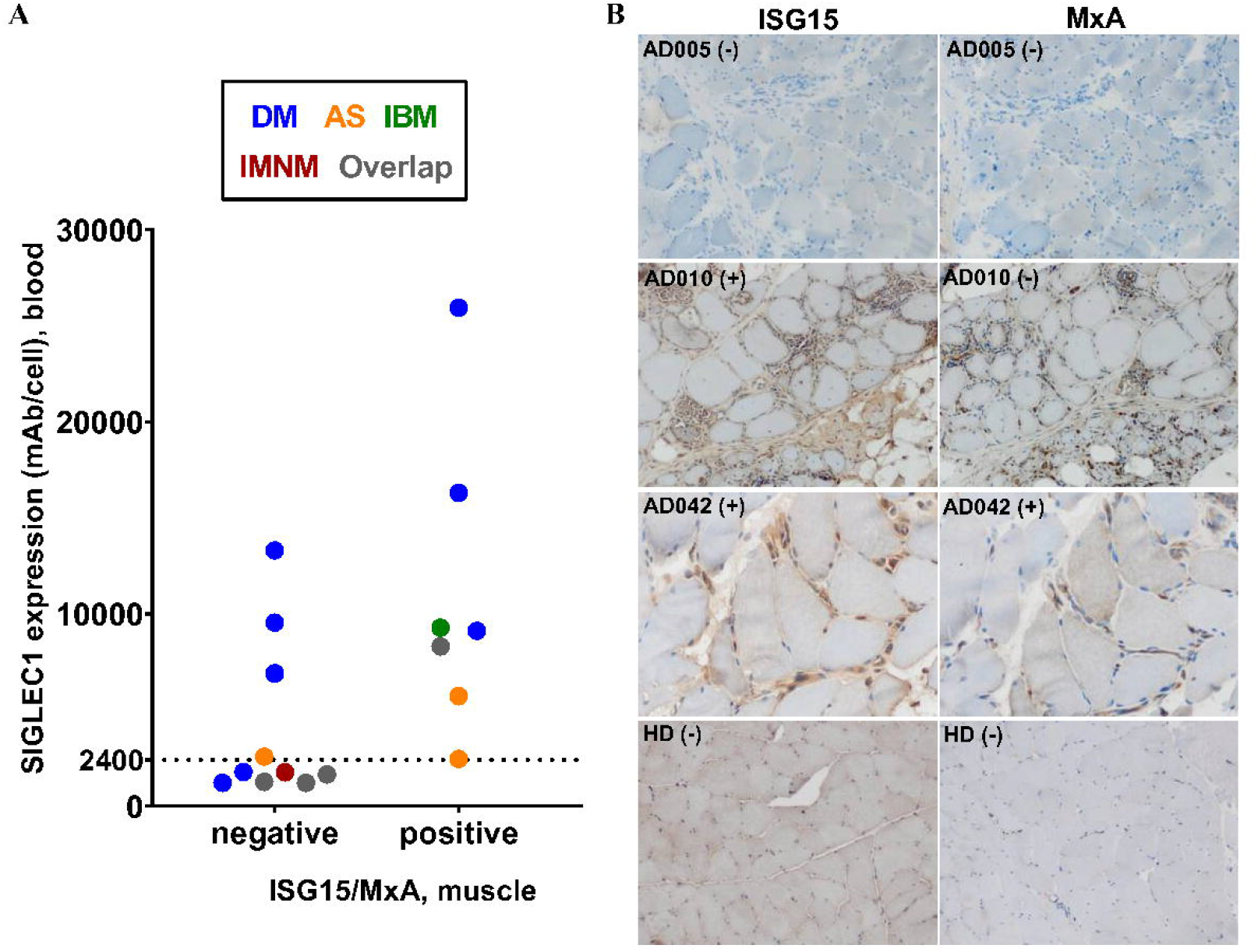
Immunohistochemical staining of type I interferon-inducible proteins (ISG15/ MxA) in muscle tissue. A) Comparison of SIGLEC1 expression in blood with negative or positive ISG15 and/or MxA status in immunohistochemical muscle biopsy staining (n=17). Colors represent patients with the specified subgroups of IIM and overlap (see legend box). B) ISG15 and MxA status of three patients and one healthy donor (HD) are shown. Patient AD005 (adult DM, SIGLEC1 in blood 13301 mAb/cell) had negative (-) stains. Patient AD010 (adult IBM, SIGLEC1 in blood 9281 mAb/cell) stained clearly positive (+) for MxA on capillaries and macrophages; regarding the ISG15 status, some macrophages were positive, but myofibers were not. Patient AD042 (adult DM, SIGLEC1 in blood 16295 mAb/cell) was ISG15-positive (more densely stained on sarcolemma than on sarcoplasm) and MxA-positive.

## Discussion

We found that SIGLEC1 expression on monocytes correlated with disease activity in patients with juvenile and adult dermatomyositis. Clinically meaningful improvement under therapy was associated with a significant decrease in SIGLEC1 expression. These results are in line with studies that analyzed type I interferon transcripts,[13–19] but with the advantage of being easily assessable by flow cytometry. Very recently, Lerkvaleekul et al.[43] published data of SIGLEC1 in 21 patients with newly diagnosed juvenile dermatomyositis. They found, that SIGLEC1 expression correlated to disease activity and was superior to predict treatment response compared to an interferon-stimulated gene score consisting of five genes. Our results validate the findings in juvenile dermatomyositis and by that underline the potential of SIGLEC1 in clinical routine diagnostics.

In our study, SIGLEC1 could distinguish between active and inactive disease in adult and juvenile dermatomyositis patients with a large area under the curve. In this context, published results in a preprint article and comment suggest the utility of direct assessment of interferon by using a highly sensitive interferon-alpha single-molecule array (SIMOA) digital enzyme-linked immunosorbent assay (ELISA) in adult and juvenile dermatomyositis.[44,45] A direct comparison of these two type I interferon biomarkers would be highly interesting, as they are both candidates for routine clinical diagnostics.

In the present study, expression of SIGLEC1 – which is mostly type I interferon-regulated[34,46] – was lower in antisynthetase syndrome than in dermatomyositis. This finding is in line with preprint published results of interferon-alpha in dermatomyositis and antisynthetase syndrome using SIMOA technology.[44] It has been proposed that type II interferons might play a more prominent role in the etiopathogenesis of antisynthetase syndrome.[39,47] However, Reed et al. found high interferon scores (including IP-10, I-TAC and MCP-1) in patients with antisynthetase syndrome.[48] We also identified patients with high expression of SIGLEC1 indicating a type I interferon response. This interesting finding warrants further investigation.

Two of our nine patients with inclusion body myositis exhibited high expression of SIGLEC1: both were positive for anti-Ro and one was also positive for anti-U1RNP autoantibodies, which are known to induce type I interferons.[49] One of these anti-U1RNP^+^ inclusion body myositis patients was also analyzed for MxA/ISG15 in muscle tissue and showed an unusual positive staining on myofibers (cf. patient AD010). Data on the role of interferons in inclusion body myositis are inconsistent and need to be clarified.[39,50,51] The detection of an activated type I interferon system might have important treatment implications in this debilitating chronic disease.

We did not find evidence of an upregulation of SIGLEC1 expression in immune-mediated necrotizing myopathy, which is in line with other current studies analyzing type I interferon-regulated transcripts (in muscle) or interferon-alpha by SIMOA technology (in blood).[39,44]

Few studies have explored differences in interferon signatures in blood according to myositis-specific autoantibody status.[48] We found the highest SIGLEC1 expression in anti-MDA5^+^ dermatomyositis patients, but could not detect a specific association of SIGLEC1 to certain myositis-specific autoantibodies in dermatomyositis otherwise. Our findings underline, that the autoantibody status alone is not predictive for the detection of a type I interferon activation. Thus, the assessment of myositis-specific autoantibodies in conjunction with the assessment of type I interferon activity might be useful for patient stratification.

The current study has strengths and limitations. A strength is that we could demonstrate that SIGLEC1, as an implemented routine biomarker, was able to validate findings from other studies investigating type I interferon signature with the advantage of being easily assessable by flow cytometry. A limitation is that muscle strength scores equivalent to the CMAS were not available for adult patients. However, the PGA is a commonly used instrument that reflects disease activity in idiopathic inflammatory myopathies.[36] Secondly, the PGA was performed retrospectively in most cases. Since the missing scores were determined by two experienced rheumatologists with access to all clinical data for each visit but blinded to SIGLEC1 expression, we are confident that we were able to reduce a potential bias.

In conclusion, analysis of SIGLEC1 expression by flow cytometry enables the individual assessment of type I interferon activity in idiopathic inflammatory myopathies. Present and published data on SIGLEC1 expression in patients with interferon-associated rheumatic and musculoskeletal diseases, interferonopathies, and viral infections confirm that SIGLEC1 is an easy-to-use type I interferon biomarker and demonstrate its potential for patient stratification, disease activity monitoring and assessment of treatment response in routine clinical practice.

## Supporting information

Checklist_STARD2015

## Abbreviations

ISG15: interferon-stimulated gene 15
MxA: myxovirus resistance protein A
MDA5: melanoma differentiation-associated protein 5
NXP2: nuclear matrix protein 2
TIF1γ: transcriptional intermediary factor 1-gamma
PL7: anti-threonyl-tRNA synthetase
PL12: alanyl-tRNA-synthetase
Jo1: anti-histidyl tRNA synthetase
SRP: signal recognition particle
HMGCR: 3-hydroxy-3-methylglutaryl-coenzyme A reductase
COVID-19: coronavirus disease 2019
EULAR/ACR: European League Against Rheumatism/American College of Rheumatology

## Notes

### Competing Interest Statement

The authors have declared no competing interest.

### Funding Statement

Funded by the Deutsche Forschungsgemeinschaft (DFG, German Research Foundation).

### Author Declarations

Prior approval for this study was obtained from the local ethics committee of the Charite - Universitaetsmedizin Berlin (application number: EA2/094/19). Informed consent was not necessary for this retrospective study.

