## Supplemental_figures_tables_text for "SIGLEC1 enables straightforward assessment of type I interferon activity in inflammatory myopathies"

**Supplemental material**

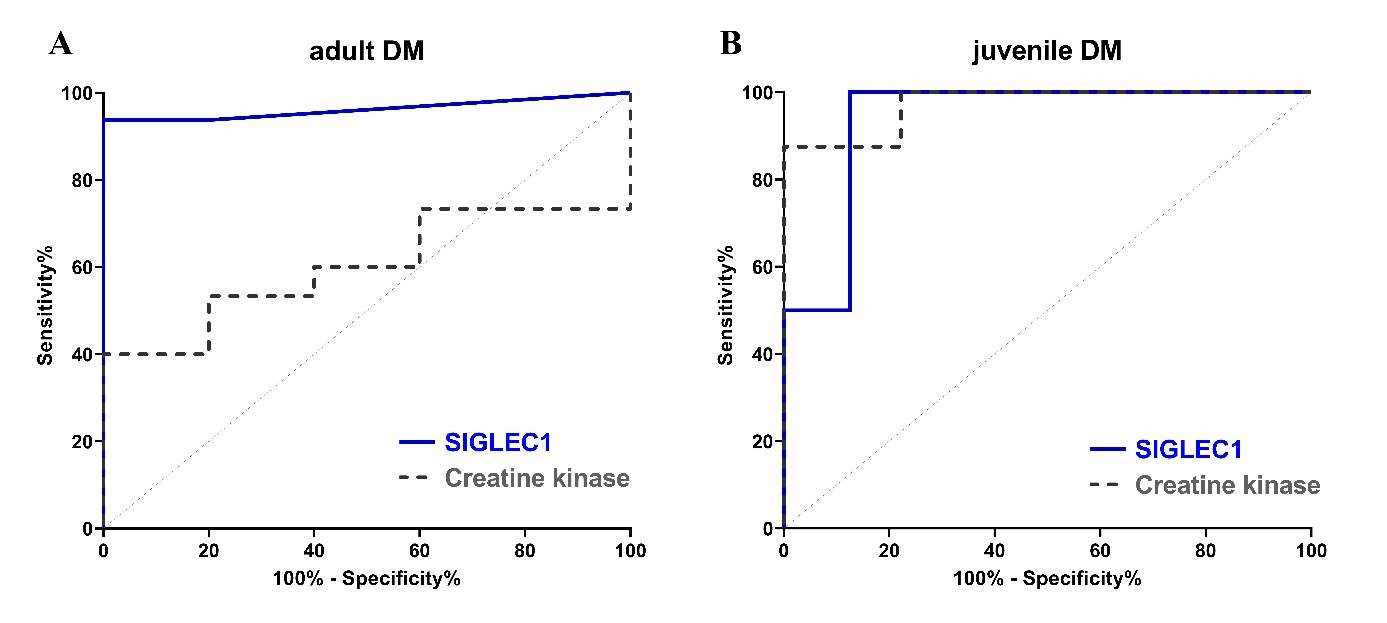

**eFigure 1 Receiver operating characteristic curves for SIGLEC1 and creatine kinase in dermatomyositis (DM) patients.** The curves show the ability of each biomarker to distinguish between patients with PGA≥5 (moderate to severe disease activity) and PGA<5 (no to moderate disease activity), in **(A)** adult DM patients (n=21); SIGLEC1: AUC=0.96, p=0.002; creatine kinase: AUC=0.60, p=0.513 and **(B)** juvenile DM patients (n=17); SIGLEC1: AUC=0.97, p=0.001; creatine kinase: AUC=0.94, p=0.003

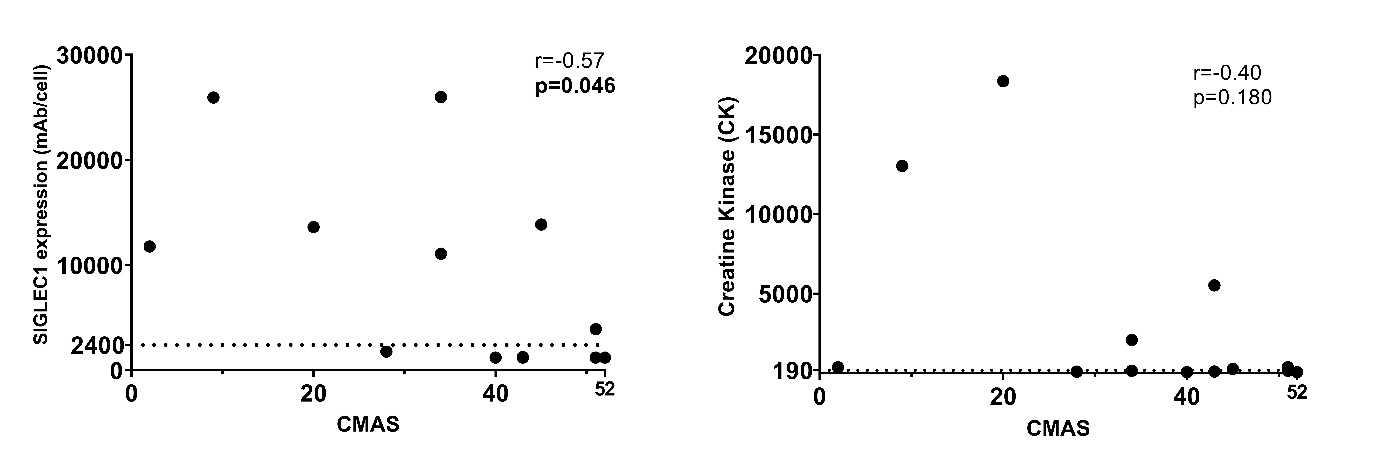

**eFigure2 Correlation between biomarkers and muscle strength scores (CMAS) in juvenile dermatomyositis patients at assessment visit 1** **(VC1)** (A) SIGLEC1 versus CMAS (n=13) and (B) creatine kinase versus CMAS (n=13); Asterisks (*) represent significant results (p<0.05). Spearman’s rank test was used for both analyses; VC1, first visit with assessment of SIGLEC1 and CMAS

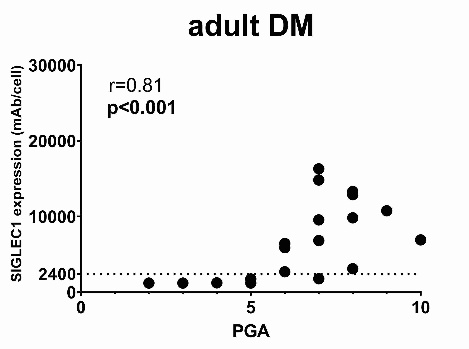

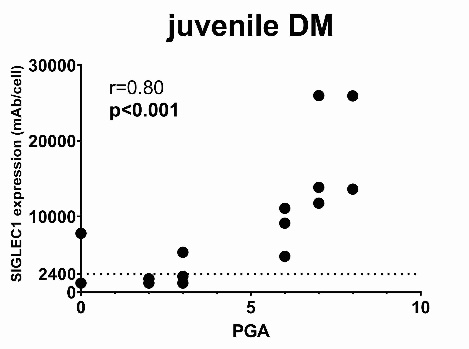

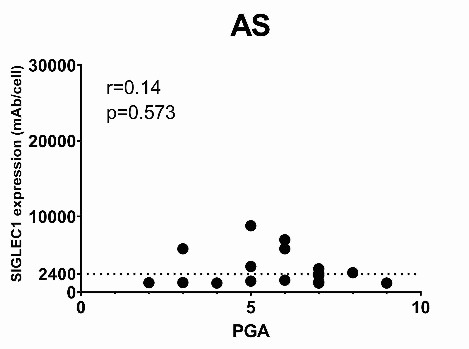

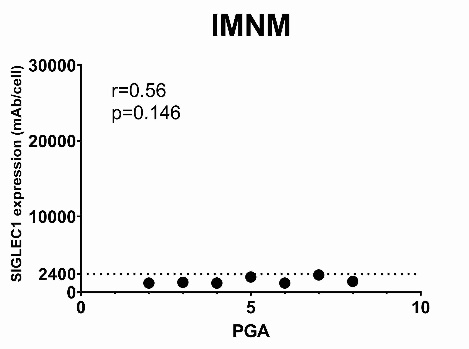

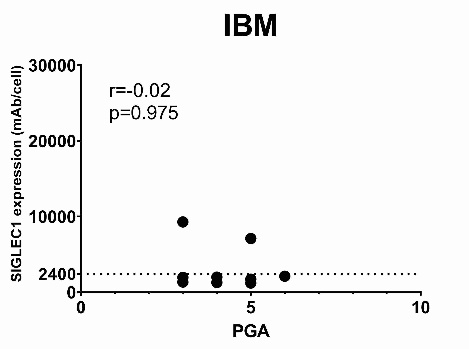

*****

*****

**eFigure3** **Correlation between SIGLEC1 expression on monocytes and Physician Global Assessment (PGA)** at first visit for the various IIM subgroups. Asterisks (*) represent significant results (p<0.05). Spearman’s rank test was used for all analyses.

| **Patient ID** | **Clinical diagnosis** | **Myositis-specific antibodies (MSA)** | **Myositis-associated antibodies (MAA)** | **SIGLEC1, mAb/cell (blood)** | **MxA status (muscle)** | **ISG15 status (muscle)** |
| --- | --- | --- | --- | --- | --- | --- |
| AD029 | AS | Jo1 | Ro52 | 5721 | 0 | 1 |
| AD031 | AS | Jo1 | Ro52 | 2568 | 0 | 0 |
| AD030 | AS | Jo1 | Ro52, PM-Scl100 | 2449 | 0 | 1 |
| AD039 | DM (adult) | Mi2 | Ro52 | 1769 | 0 | 0 |
| AD042 | DM (adult) | NXP2 | - | 16295 | 1 | 1 |
| AD005 | DM (adult) | - | Ro52 | 13301 | 0 | 0 |
| AD041 | DM (adult) | TIF1γ | - | 9545 | 0 | 0 |
| AD067 | DM (adult) | MDA5 | PM-Scl100 | 1200 | 0 | 0 |
| AD065 | DM (adult) | MDA5 | - | 6901 | 0 | 0 |
| PAE002 | DM (juvenile) | PL7 | - | 9114 | 0 | 1 |
| PAE001 | DM (juvenile) | - | - | 25931 | 1 | 1 |
| AD010 | IBM | - | Ro52, Ku, U1RNP | 9281 | 1 | 0 |
| AD004 | Overlap | - | Ku | 8305 | 0 | 1 |
| AD072 | IMNM | HMGCR | - | 1752 | 0 | 0 |
| AD008 | Overlap | - | Ro52, Ku, PM-Scl75, PM-Scl100 | 1645 | 0 | 0 |
| AD047 | Overlap | SRP | - | 1260 | 0 | 0 |
| AD064 | Overlap | - | Ku | 1200 | 0 | 0 |

**eTable1** **Subgroup analysis of SIGLEC1 expression (in blood) versus MxA and ISG15 status** (in immunohistochemical muscle biopsy stains) (n=17). MxA/ISG15 status: 0 = negative, 1 = positive

**eFigure4** **Longitudinal graphs of SIGLEC1 vs. CMAS and creatine kinase vs. CMAS** for all juvenile DM patients with at least two visits; PGA, Physician Global Assessment; CMAS, Childhood Myositis Assessment Scale

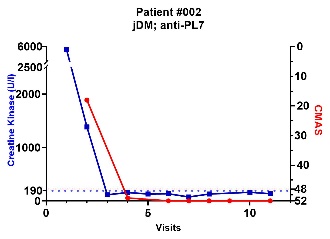

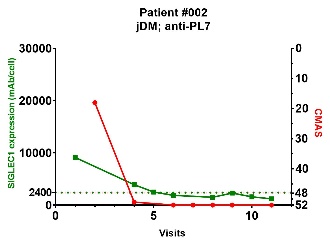

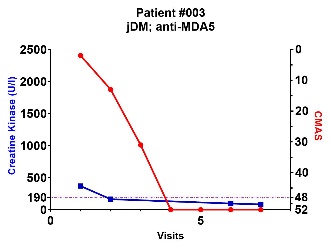

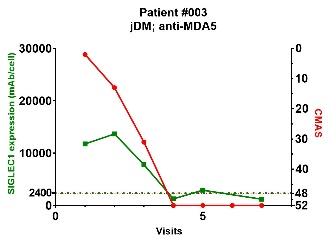

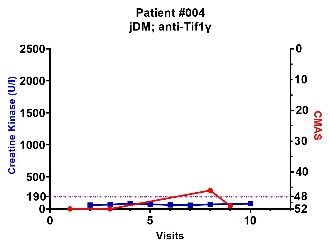

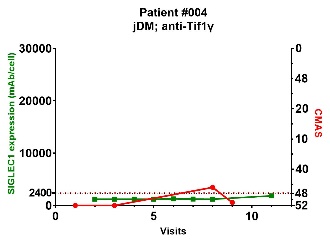

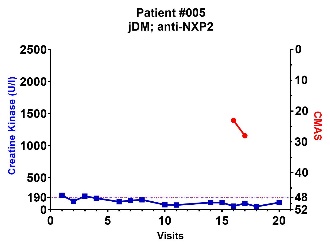

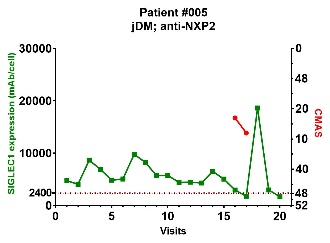

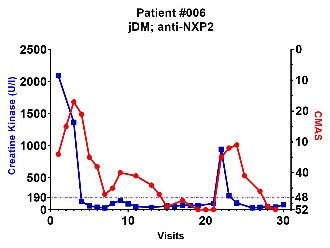

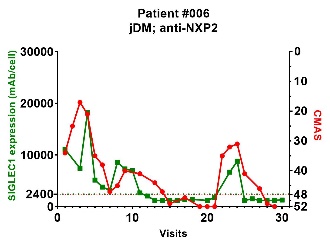

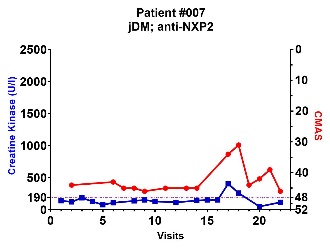

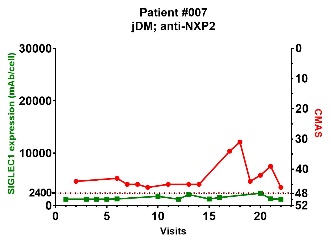

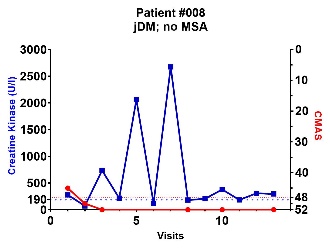

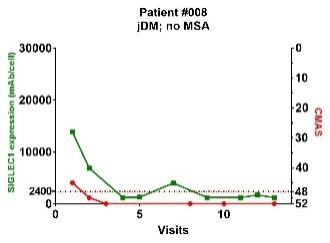

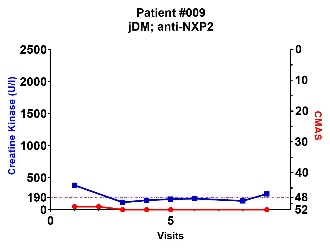

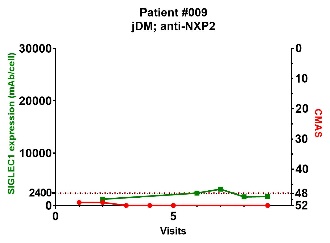

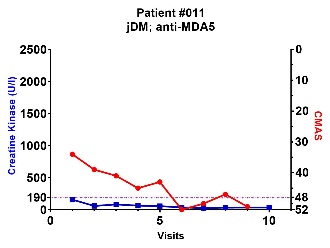

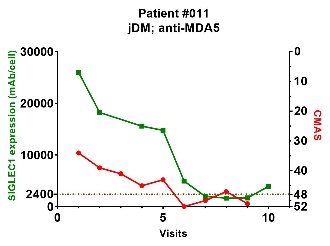

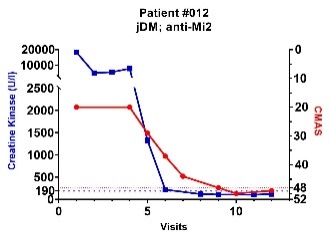

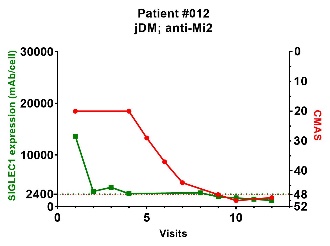

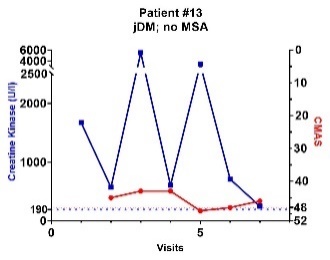

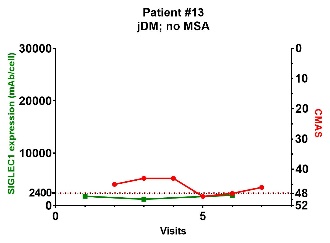

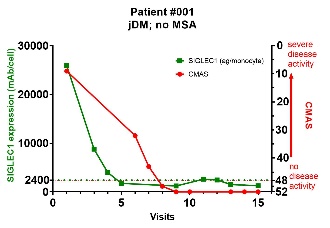

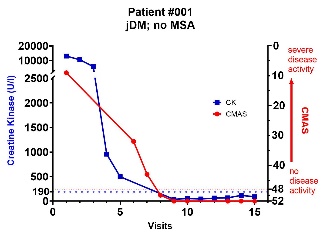

**eFigure5** **Longitudinal graphs of SIGLEC1 vs. PGA and creatine kinase vs. PGA** for all adult DM patients with at least two visits; PGA, Physician Global Assessment; CMAS, Childhood Myositis Assessment Scale

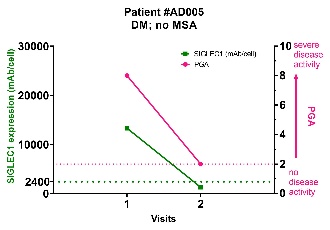

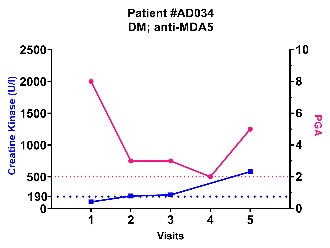

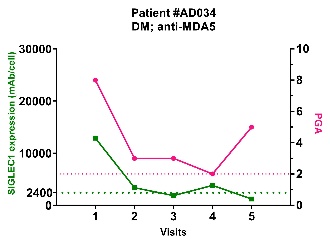

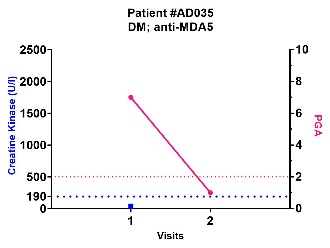

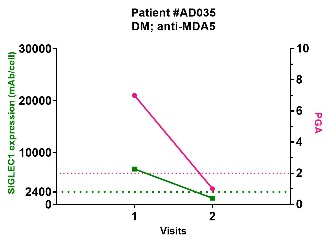

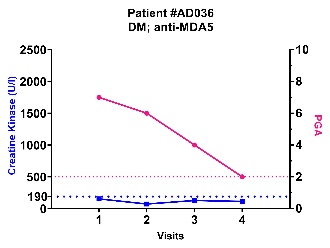

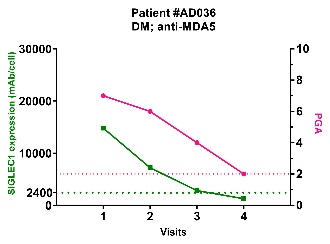

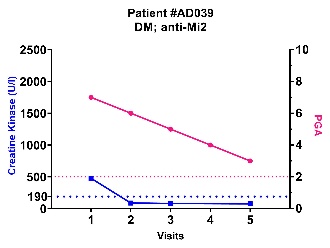

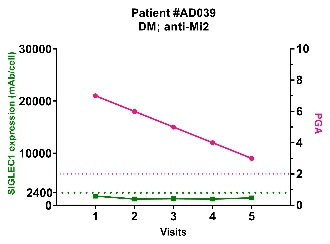

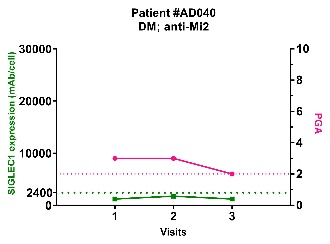

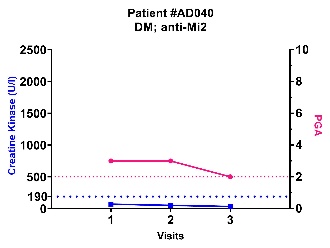

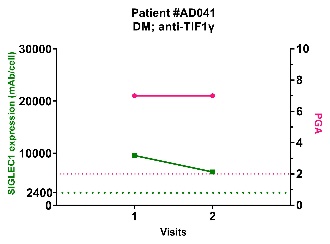

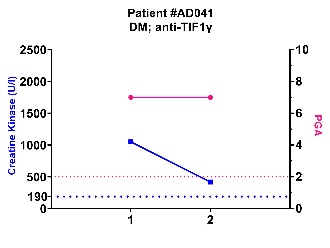

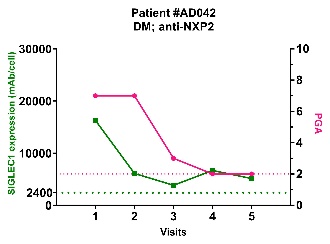

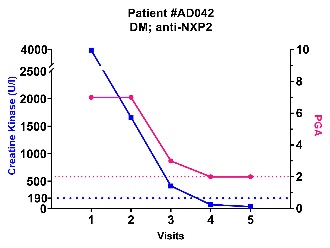

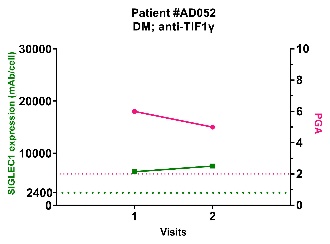

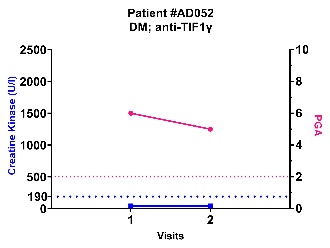

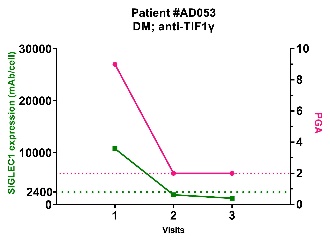

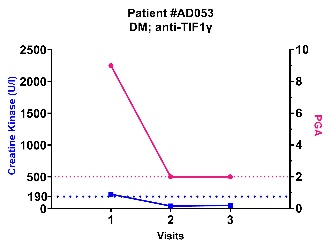

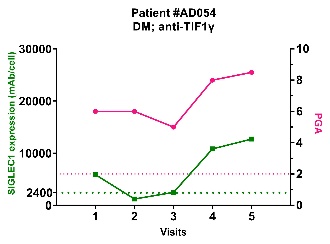

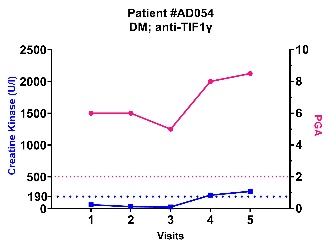

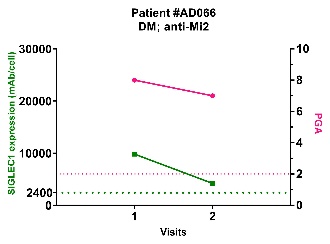

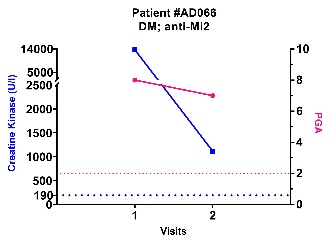

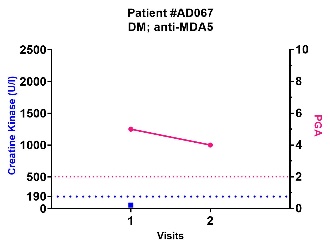

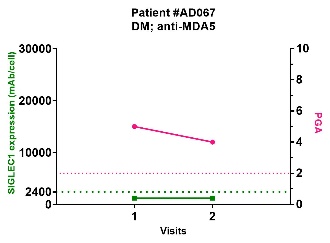

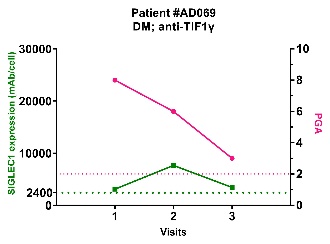

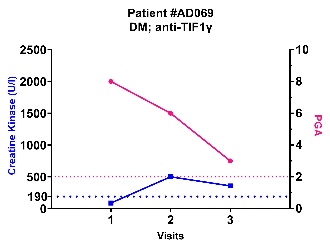

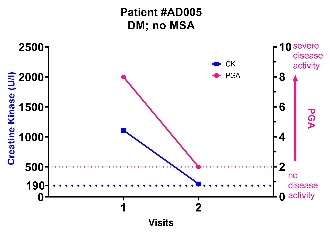

**eFigure6** **Visualization of the results of longitudinal data analysis** (see Table 2) of A) SIGLEC1 vs. CMAS for juvenile DM patients (absolute values); B) Creatine kinase (CK) vs. CMAS for juvenile DM patients (absolute values); C) SIGLEC1 vs. PGA for adult DM patients (absolute values); D) Creatine kinase (CK) vs. PGA for adult DM patients (absolute values)

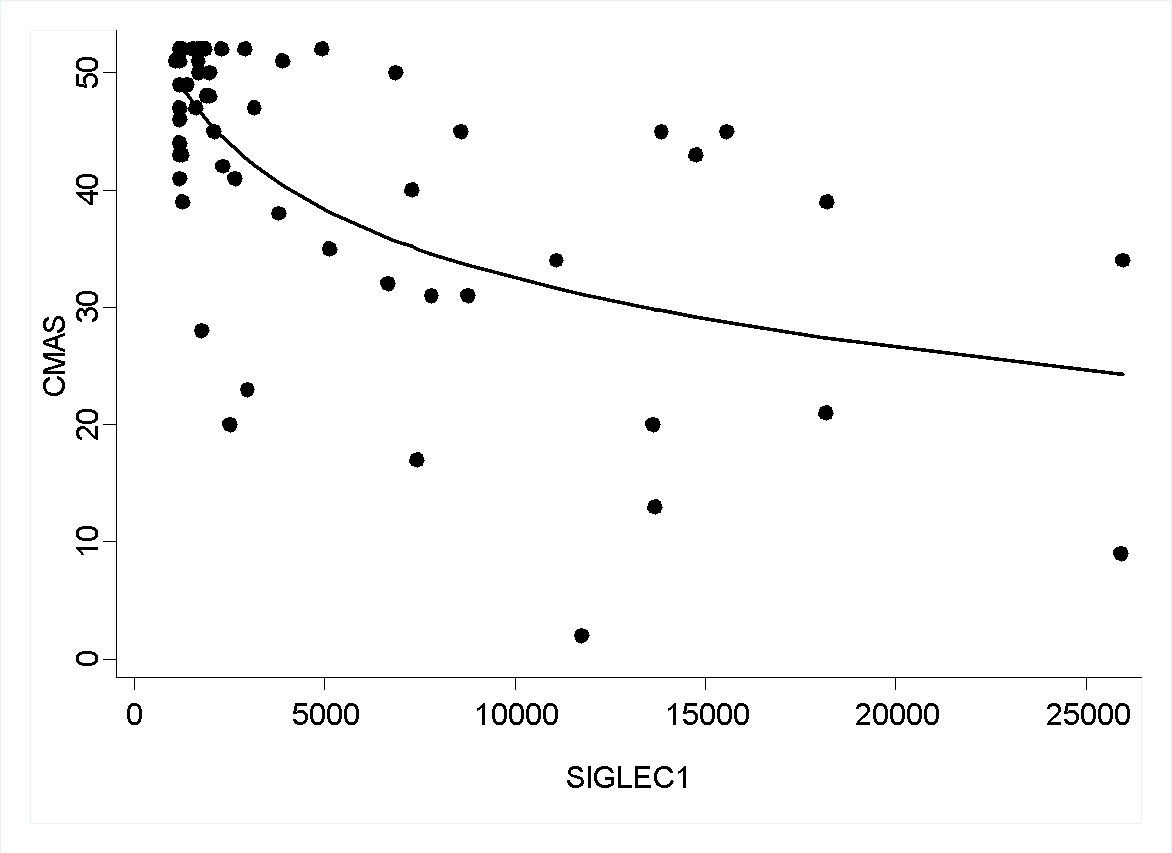

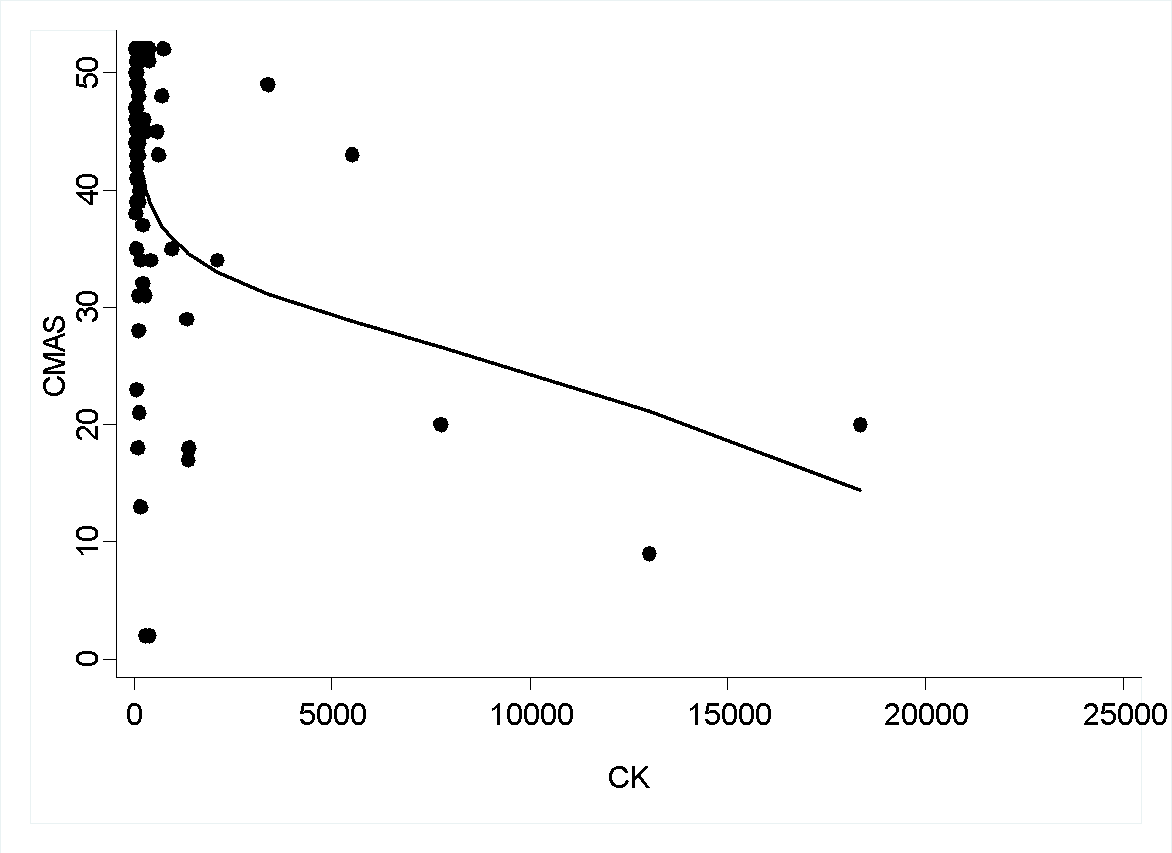

**A**

**B**

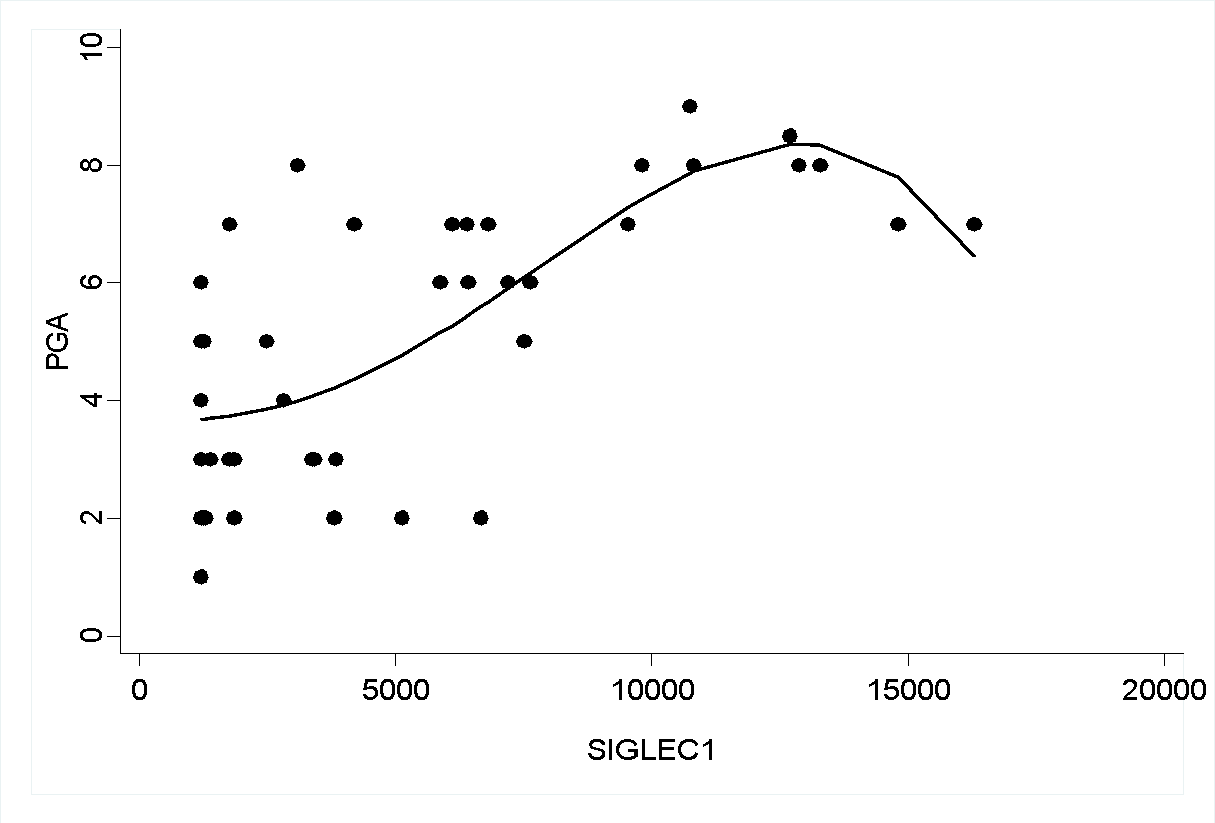

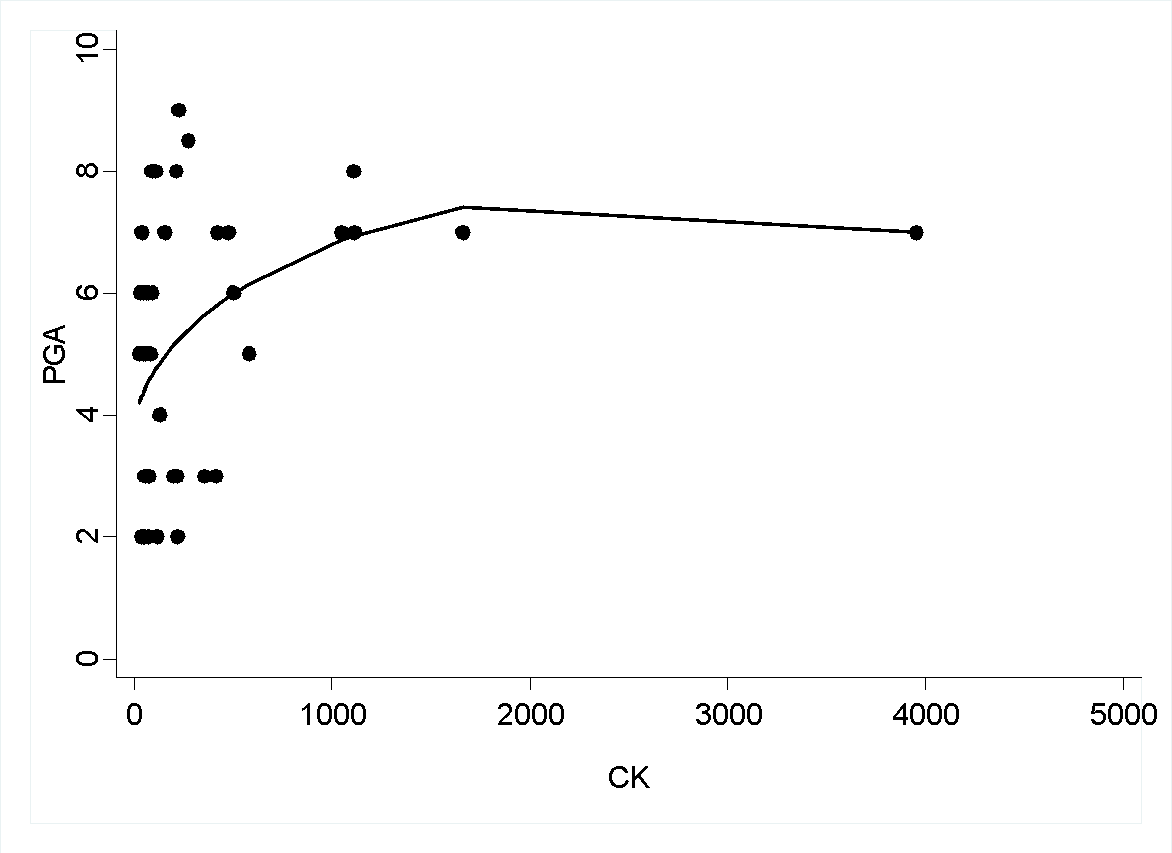

**D**

**C**

**eText1: Laboratory measurement of SIGLEC1**

SIGLEC1 expression was determined by flow cytometry using a highly standardized quantitative assay. Briefly, 25µl of EDTA-anticoagulated whole blood was incubated with 10 µl of mouse-anti-human antibody cocktail containing phycoerythrin (PE)-labeled anti-CD169 monoclonal antibody (mAb) (labeled at a fluorochrome/protein ratio of 1:1), allophycocyanin (APC)-labeled anti-CD14 mAb and KromeOrange-labeled anti-CD45 mAb for 15 min at room temperature (RT) in the dark (all antibodies from Beckman Coulter, Krefeld, Germany). Red blood cells were then lysed by adding 500µl of Versa-Lysis solution (Beckman Coulter) to each reaction tube. After incubation for 30 min at RT in the dark,

samples were centrifuged for 5 min at 200 x g at RT. Samples were then washed once with 1000 µl PBS containing 2% fetal calf serum (FCS) and centrifuged again for 5 min at 200 x g at RT. Stained samples were acquired on a 10-color flow cytometer (Navios, Beckman Coulter) and analyzed using the Navios software. During each analytical run, QuantiBRITE™ PE tubes (BD Biosciences) were acquired to convert the fluorescent channel 2 (FL2) mean fluorescent intensity (MFI) signals on CD14^+^ monocytes to monoclonal antibodies bound per cell (mAb/cell) values. FL2 MFI values and absolute values for PE molecules (as specified by the manufacturer) for each QuantiBRITE™ bead population were used to perform linear least square regression analysis to determine the best calibration value, which then was used to convert the FL2 MFI values of monocytes in the analytical sample to the amount of PE-labeled CD169 mAb bound per monocyte (mAb/cell).

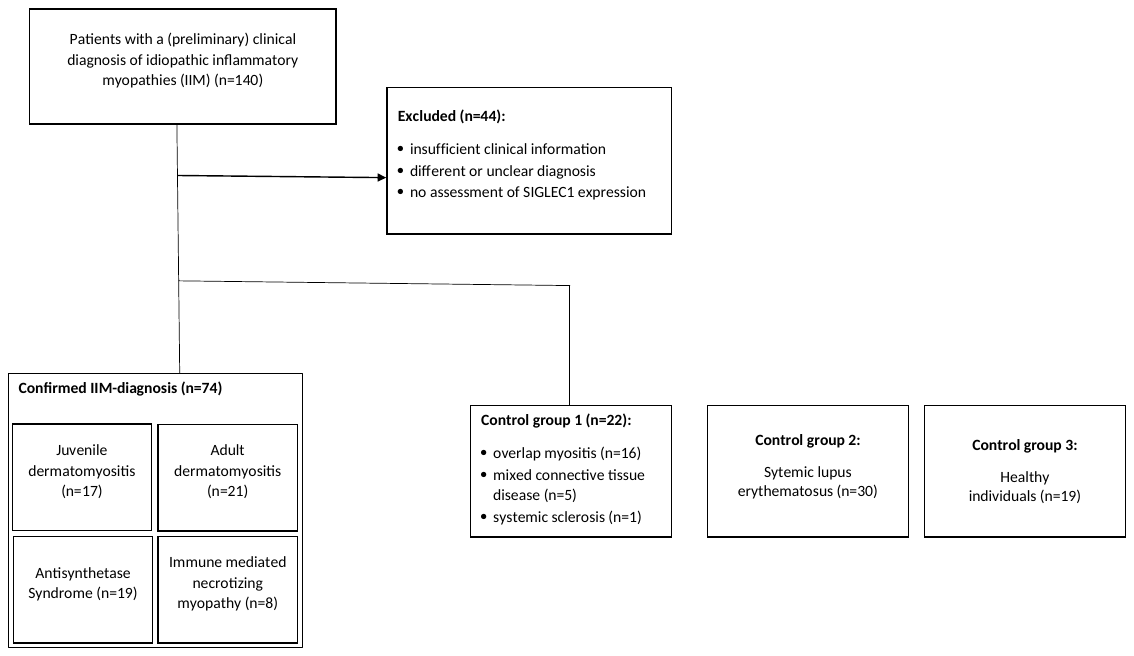

**eFigure 7** Flow diagram of study participants
